# Supply Chain Vulnerabilities in First-Line Treatments for Sexually Transmitted Infections: Implications for U.S. Public Health Preparedness

**DOI:** 10.64898/2026.05.06.26352546

**Authors:** Coby Y Garcia, William Leung, Amelia Shirley, Ian Zhao, Lao-Tzu Allan-Blitz

**Author notes:** POST-PUB CORRESPONDING AUTHOR: Lao-Tzu Allan-Blitz, MD, MPH, Division of Infectious Diseases, Department of Medicine University of California Los Angeles, 10833 Le Conte Ave, Los Angeles, CA 90095. PRE-PUB CORRESPONDING AUTHOR, Coby Y. Garcia, 1864 Singleton Street Indianapolis, IN, USA.

## Abstract

**Objectives:** To evaluate supply-chain vulnerabilities affecting medications essential for treating sexually transmitted infection in the United States and identify disruption mechanisms that may predispose these therapies to shortages.

**Methods:** We conducted a qualitative, structured supply-chain vulnerability assessment of first-line medications for five priority sexually transmitted pathogens recommended by the Centers for Disease Control and Prevention and the World Health Organization: azithromycin, doxycycline, ceftriaxone, benzathine penicillin G, metronidazole, tinidazole, acyclovir, and cefixime. Using a predefined framework derived from pharmaceutical supply-chain disruption literature, we evaluated 13 disruption categories spanning raw material sourcing, active pharmaceutical ingredient production, manufacturing, distribution, market dynamics, information systems, and post-distribution loss mechanisms. Each category was assessed using four binary indicators and classified as relevant when at least two criteria were satisfied.

**Results:** Multiple disruption domains applied across the drug set. Recurrent vulnerabilities included geographically concentrated active pharmaceutical ingredient production, limited manufacturing redundancy in low-margin generic markets, manufacturing constraints affecting sterile injectable products, reliance on consolidated distribution networks, and susceptibility to demand surges and information-system disruptions. All eight drugs exhibited at least one regulatory or market signal consistent with potential supply vulnerability, including documented shortages, product discontinuations, or limited manufacturer participation.

**Conclusions:** Supply-chain vulnerabilities were identified across multiple first-line sexually transmitted infection therapies, indicating that disruption risk is not confined to a single drug. There is a need for policy interventions to strengthen supply-chain resilience, including diversification of active pharmaceutical ingredient sourcing and distribution networks, as well as incentives for sustainable generic production.

## INTRODUCTION

Access to effective antimicrobial therapies is essential for treating sexually transmitted infections (STIs). The consequences of untreated STIs include pelvic inflammatory disease, infertility, obstetric complications, neonatal death, adverse birth outcomes, and an increased risk for the transmission of HIV ^6^. In June 2023, Pfizer warned that it would soon run out of benzathine penicillin G ^7^. Because Pfizer was the only manufacturer supplying the medication to the United States (U.S.), and because penicillin is the only recommended treatment for syphilis during pregnancy, the shortage created a critical challenge for preventing congenital syphilis ^1,8^. The loss of effective treatment coincided with a resurgence of syphilis and congenital syphilis in the U.S.; since 2012, the number of congenital syphilis cases has increased by more than 1,000%^9^. The shortage of benzathine penicillin G drew attention to structural weaknesses in the pharmaceutical supply chain with direct consequences for patient care and public health.

Drug shortages arise from vulnerabilities across pharmaceutical supply chains, including upstream sourcing, manufacturing concentration, distribution, and market dynamics ^2,3^. Although prior studies have examined drug shortages broadly, few have focused on STI treatments, many of which are generic, low-margin products with constrained manufacturing bases ^5,10,11^. Whether those drugs share vulnerabilities similar to benzathine penicillin G remains unclear.

In this study, we evaluated supply-chain vulnerabilities across eight core STI drugs recommended by the U.S. Centers for Disease Control and Prevention and the World Health Organization (WHO). Using a structured qualitative framework based on 13 pharmaceutical supply-chain disruption categories, we assessed risks spanning raw materials, manufacturing, distribution, information systems, and market behavior. By grounding our analysis in regulatory data, documented shortage histories, and established disruption mechanisms, we sought to identify which drugs used to treat priority sexually transmitted pathogens may be vulnerable to disruptions analogous to the recent U.S. penicillin shortage and to inform strategies for strengthening supply-chain resilience.

## METHODS

### Research Design

We conducted a qualitative, structured supply-chain vulnerability assessment of core medications used to treat STIs in the U.S. We applied a rule-based framework described below to evaluate whether established pharmaceutical supply-chain disruption factors met predefined criteria for relevance to each drug. We designed the study as a structured qualitative assessment designed to identify recurring categories of supply-chain vulnerability across drugs.

### Drug Selection

We focused on the five priorities STIs identified in the U.S. Sexually Transmitted Infections National Strategic Plan (2021–2025) and its addendum: *Chlamydia trachomatis, Neisseria gonorrhoeae, Treponema pallidum, Trichomonas vaginalis*, and herpes simplex virus types 1 and 2. ^12,13^.

We selected the first-line treatment options for each STI recommended by the CDC or WHO STI treatment guidelines as well as acceptable alternative first-line therapies. We excluded therapies recommended only under restricted clinical conditions (e.g., allergy-specific substitutions, salvage regimens for resistant infections, or context-dependent indications).

### Data Collection

We used a structured, multi-source data collection strategy in which each disruption category was evaluated using sources from multiple types of datasets. For peer-reviewed evidence, we searched MEDLINE and Embase for each included drug using predefined combinations of drug names and disruption-related terms. Those searches were designed to identify evidence on formulation characteristics, historical shortages, manufacturing disruptions, distribution constraints, demand surges, recalls, supplier concentration, and related supply-chain vulnerabilities. Full search terms for each drug and concept combination are provided in the Appendix.

For U.S. regulatory and product-market evidence, we reviewed several FDA databases to systematically characterize the structure and resilience of drug supply chains: 1) Drugs@FDA to identify approved new drug applications and abbreviated new drug applications^15^, 2) the FDA Drug Shortages Database to identify active and resolved shortages and manufacturer-reported causes^16^, 3) the Orange Book to assess therapeutic equivalence and approval context^17^, and 4) the National Drug Code Directory to identify listed products and marketed presentations^18^. Together, those sources enabled us to assess key dimensions of supply vulnerability, including manufacturer concentration, approval pathways, shortage history, and marketed-product availability.

For disruption categories that were not systematically indexed or consistently retrievable in bibliographic databases, such as misinformation-driven demand surges, off-label promotion, cyber incidents, trade disputes, and certain distribution shocks, we used a predefined gray-literature search strategy. Specifically, we searched Google search engine as well as specific U.S. government websites: FDA, CDC, Department of Health and Human Services, Administration for Strategic Preparedness and Response, Drug Enforcement Administration, and U.S. Customs and Border Protection when relevant. We also searched international and multilateral websites: WHO and European Medicines Agency.

We included industry and supply-chain sources such as manufacturer-issued recall notices, FDA enforcement reports, drug shortage communications, and publications from major pharmaceutical wholesalers (e.g., AmerisourceBergen, McKesson, and Cardinal Health) and pharmacy organizations (e.g., American Society of Health-System Pharmacists). Finally, we searched the following archival news databases and major news outlets when a disruption event was reported publicly but not yet represented in peer-reviewed or regulatory literature: *The New York Times*, *The Wall Street Journal*, *Reuters*, *Associated Press*, *STAT News*, and *Bloomberg*, as well as LexisNexis and Factiva. We specified the exact website domains, date ranges, and keyword strings used for those searches in the Appendix.

For all sources, we aligned search terms with the corresponding binary assessment questions. For example, searches paired each drug name with terms such as “drug shortage,” “supply chain,” “manufacturing disruption,” “recall,” “API,” “active pharmaceutical ingredient,” “demand surge,” “counterfeit,” “diversion,” “cyberattack,” “export restriction,” and “trade dispute,” as appropriate to the disruption category. We prioritized sources that documented population-level, system-level, or market-level relevance rather than isolated anecdotal events.

The Appendix contains the complete search syntax, database interfaces, websites searched, and the decision rules used to determine whether retrieved evidence satisfied a given binary criterion.

### Measures

We evaluated supply-chain vulnerability using disruption categories adapted from prior work, spanning raw material sourcing, manufacturing, distribution, market dynamics, and information systems^14^. For each disruption category, we developed four binary assessment questions designed a priori to determine whether the disruption mechanism was relevant to a given drug. A disruption category was classified as relevant when at least two questions were answered “yes” with documented supporting evidence. Two authors independently evaluated each question. Disagreements were resolved by a third author. The binary classification was used to standardize application of the framework across drugs rather than to quantify the magnitude or severity of vulnerability.

### Ethical Considerations

This study included only publicly available, de-identified databases. As such, it was exempt from institutional review.

## RESULTS

### Identification and Selection of STI Drugs

We identified 14 pharmacologic agents recommended as either first-line or alternative first-line regimens for the five priority STIs (Table 1). Of the 14 identified agents, six were excluded prior to analysis due to therapeutic redundancy (*n* = 2), niche or allergy-restricted use (*n* = 1), salvage therapy for rare resistant infections (*n* = 1), or context-specific alternatives with inconsistent representation across federal databases (*n* = 2) (Table S1). The remaining eight drugs (azithromycin, doxycycline, ceftriaxone, cefixime, benzathine penicillin G, metronidazole, tinidazole, and acyclovir) comprised the final analytic sample used for structured supply-chain vulnerability assessment (Table 2).

**Table 1.**
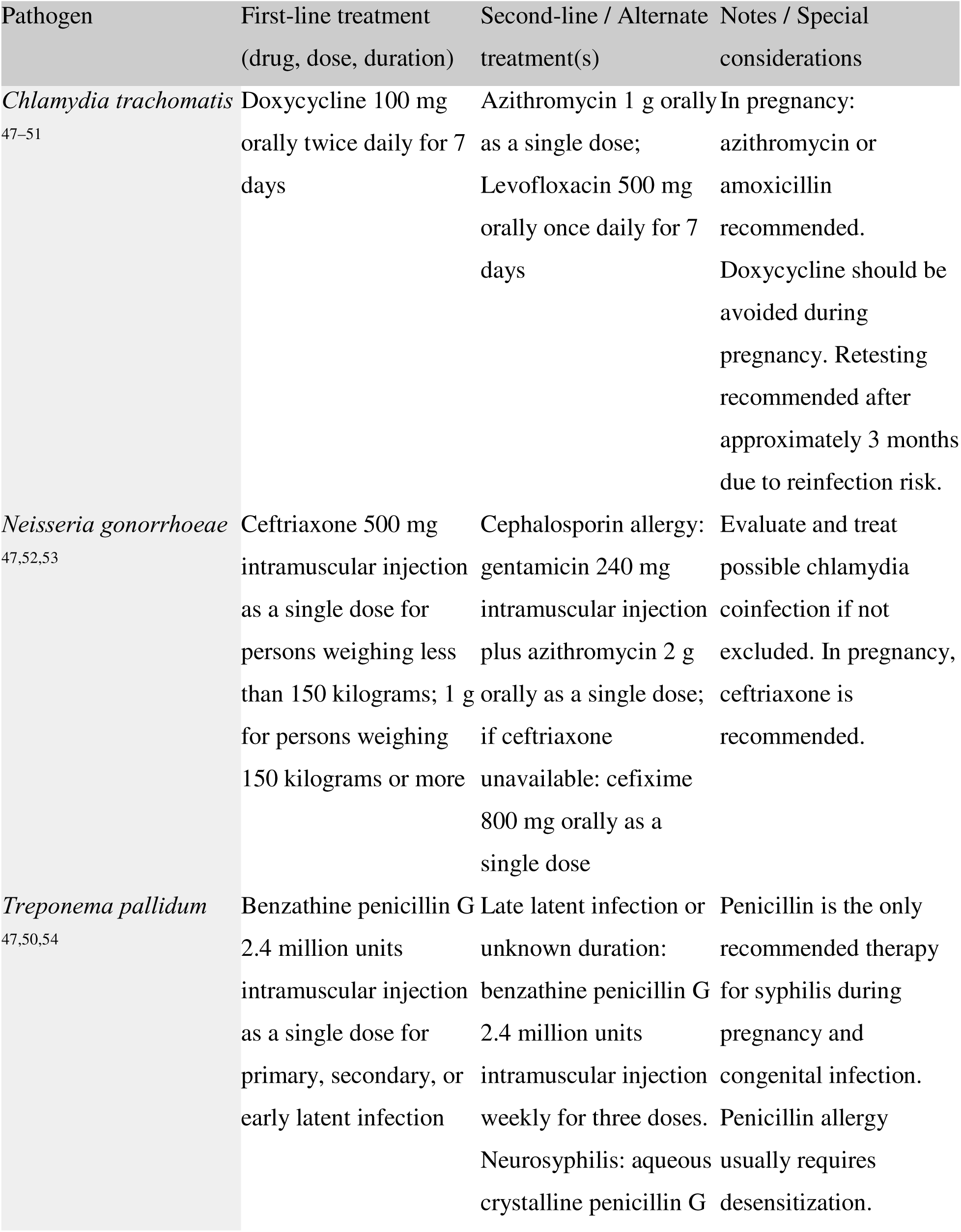
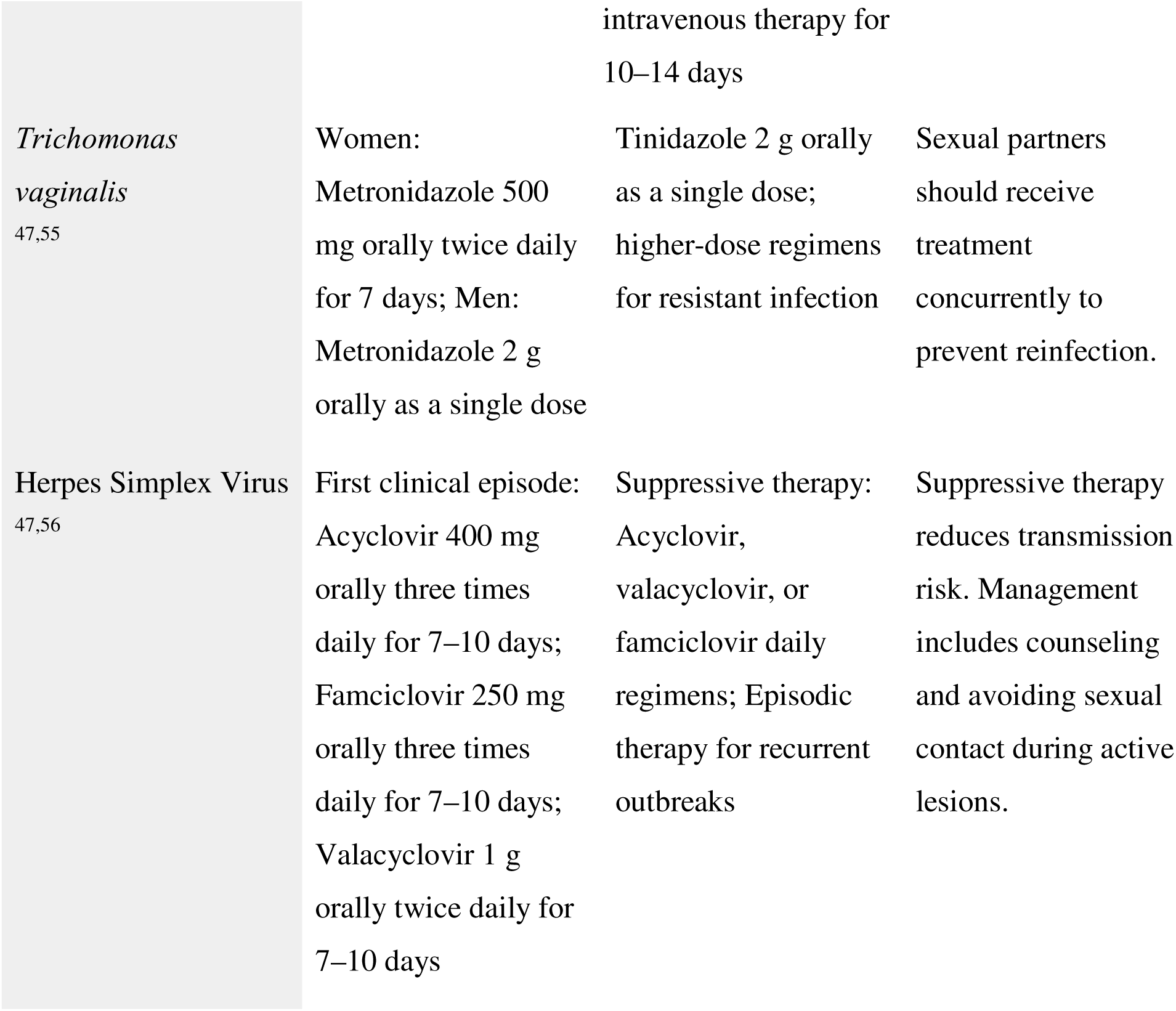
First-Line and Alternative First-Line Pharmacologic Treatments for Priority Sexually Transmitted Infections, United States, 2021–2025.

**Table 2.**
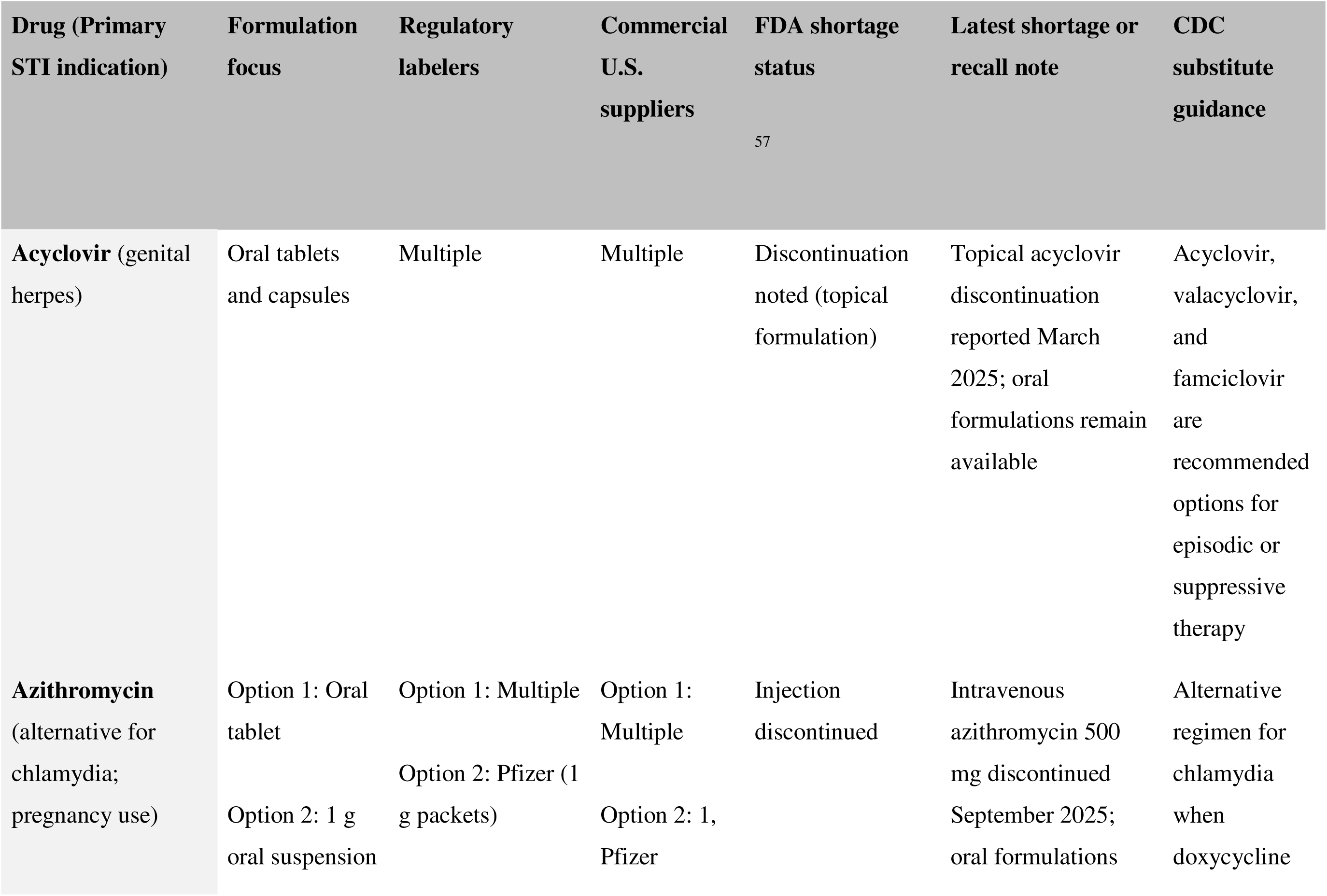
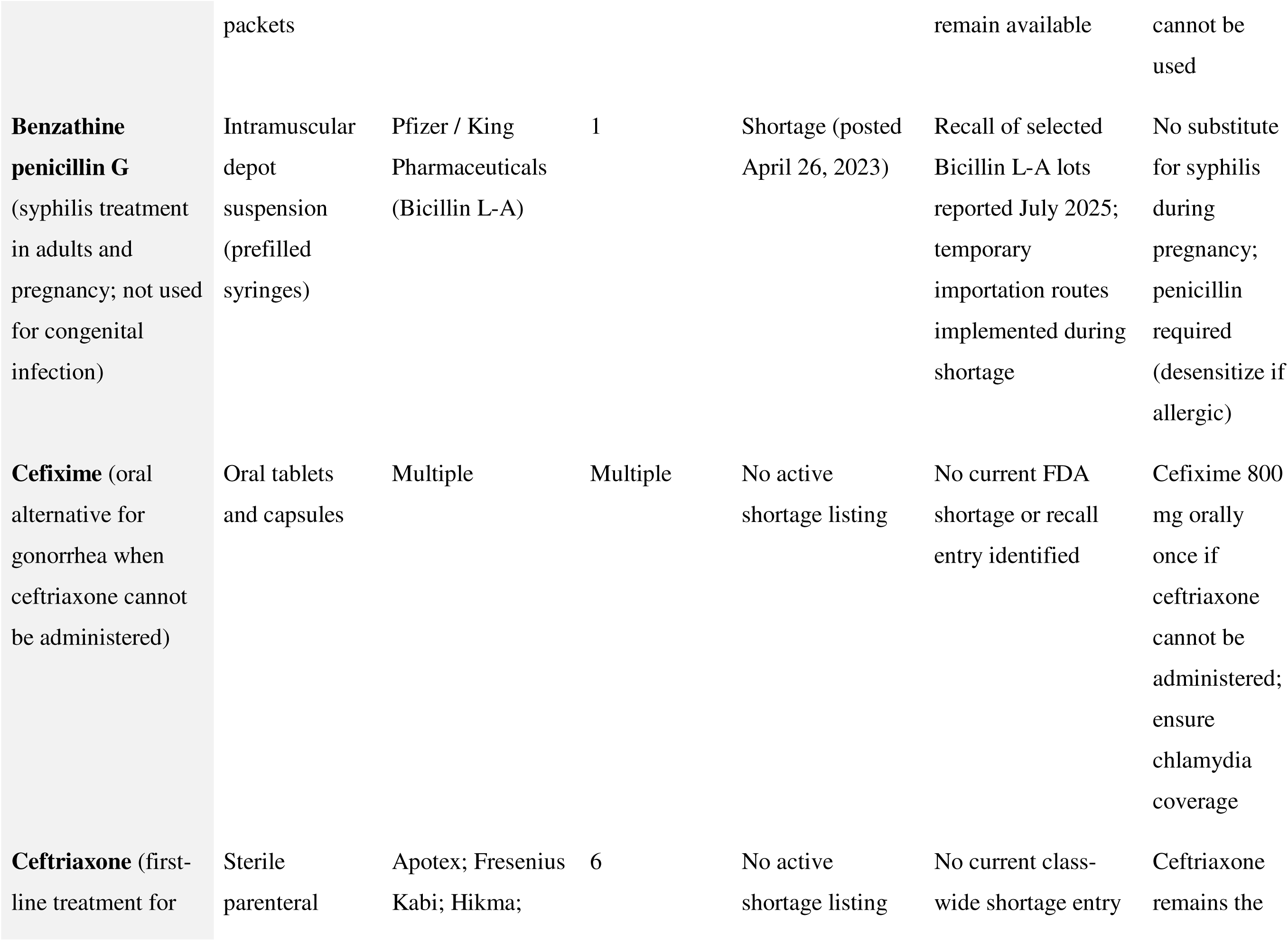
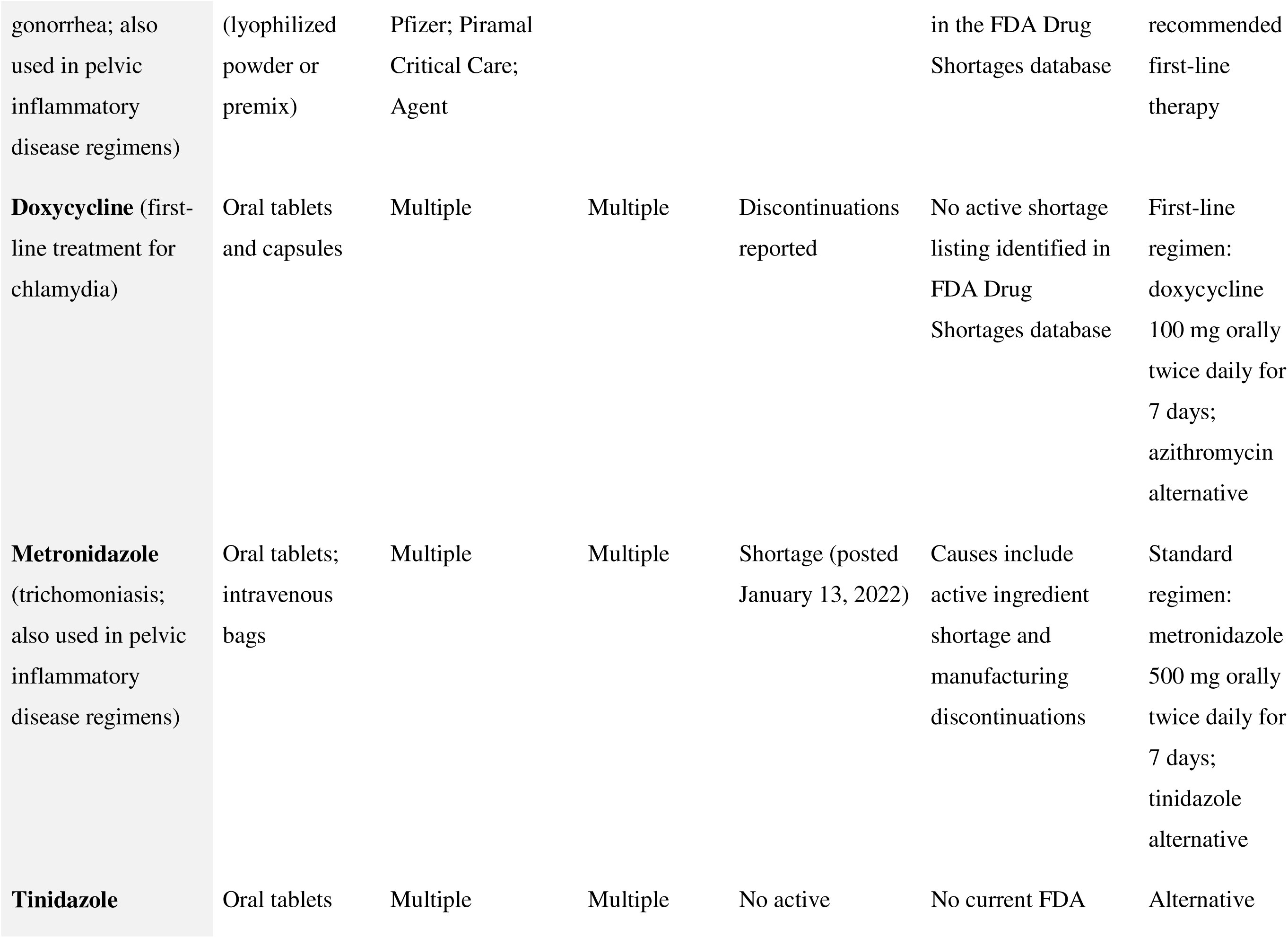
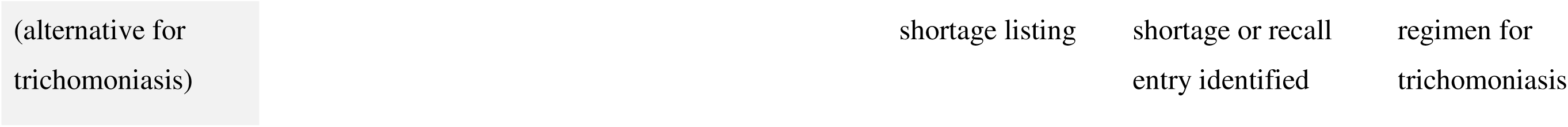
Regulatory Status, Formulation Type, Manufacturer Participation, and Shortage History of Eight STI Medications, United States, 2022–2025.

### Regulatory Status, Manufacturer Landscape, and Shortage History

Table 2 summarizes regulatory approval status, formulation type, commercially active U.S. manufacturers, and documented shortage or discontinuation history for the eight included drugs. Six drugs were oral solid-dose generics (azithromycin, doxycycline, cefixime, metronidazole, tinidazole, and acyclovir). Two drugs, ceftriaxone and benzathine penicillin G, were sterile injectable formulations requiring aseptic manufacturing and parenteral administration.

Manufacturer participation varied substantially across drugs. Oral generics were produced by multiple manufacturers, whereas benzathine penicillin G and the oral suspension packets of Azithromycin had a single primary U.S. supplier, indicating high manufacturing concentration. Ceftriaxone had six identified manufacturers, and the other six drugs had more than one U.S. supplier.

FDA Drug Shortages Database records documented prior shortages for benzathine penicillin G (shortage posted April 2023) and metronidazole (shortage posted January 2022), with additional recall and supply disruption events noted for benzathine penicillin G in 2025 (Table 3). No active or historical shortage entries were identified for cefixime, tinidazole, or acyclovir. Ceftriaxone had no active class-wide shortage listing at the time of review despite its reliance on sterile manufacturing.

**Table 3.**
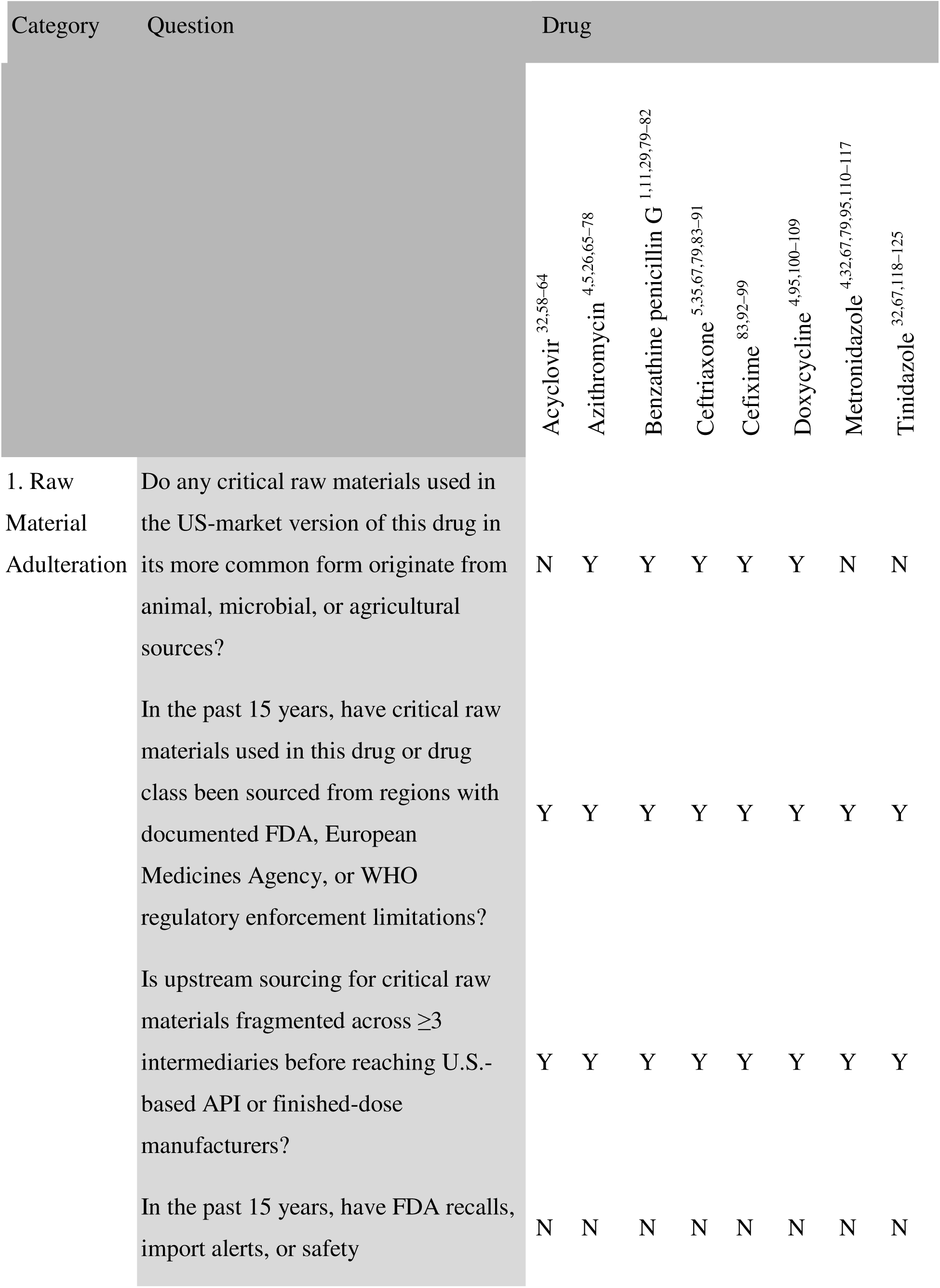
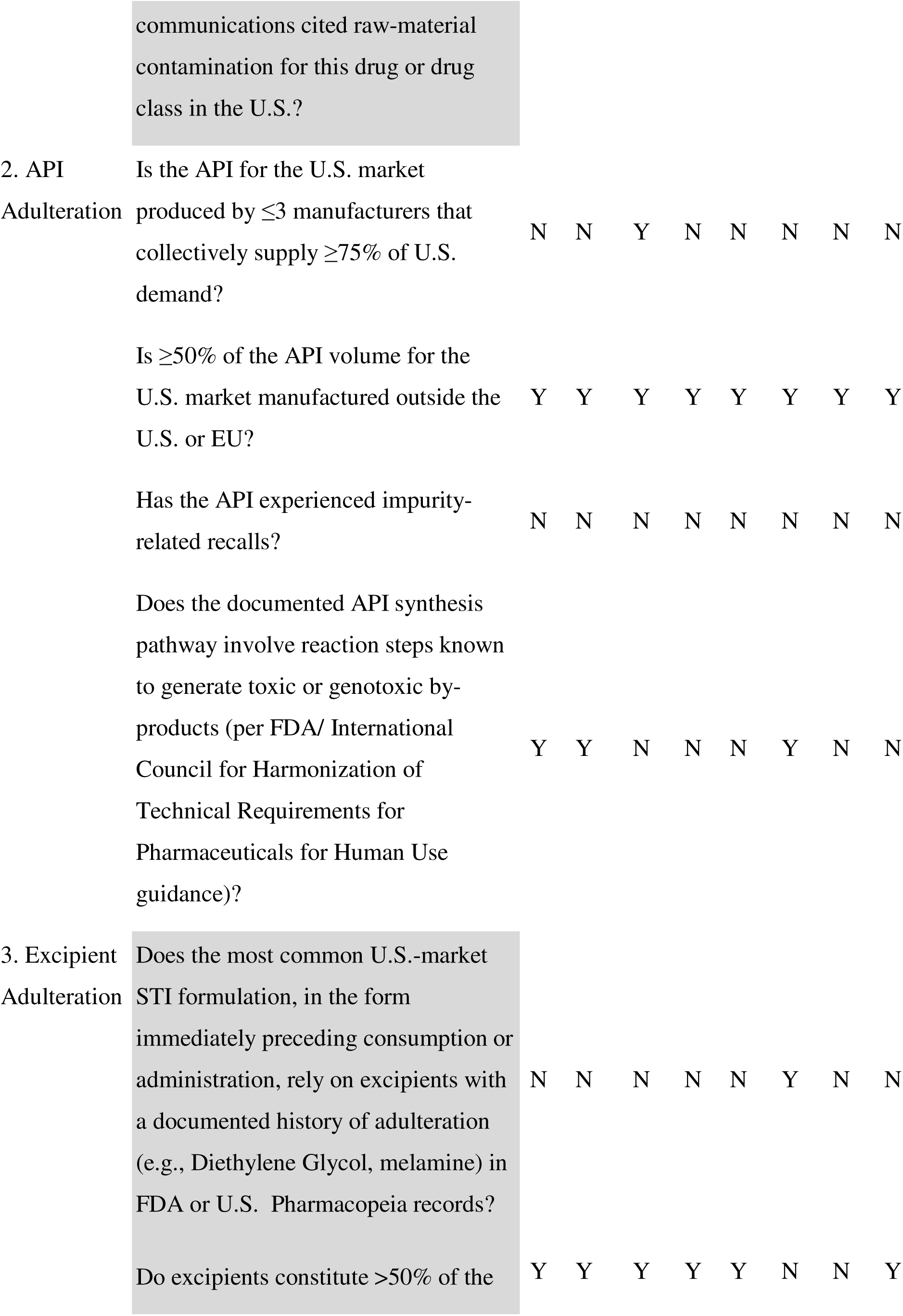
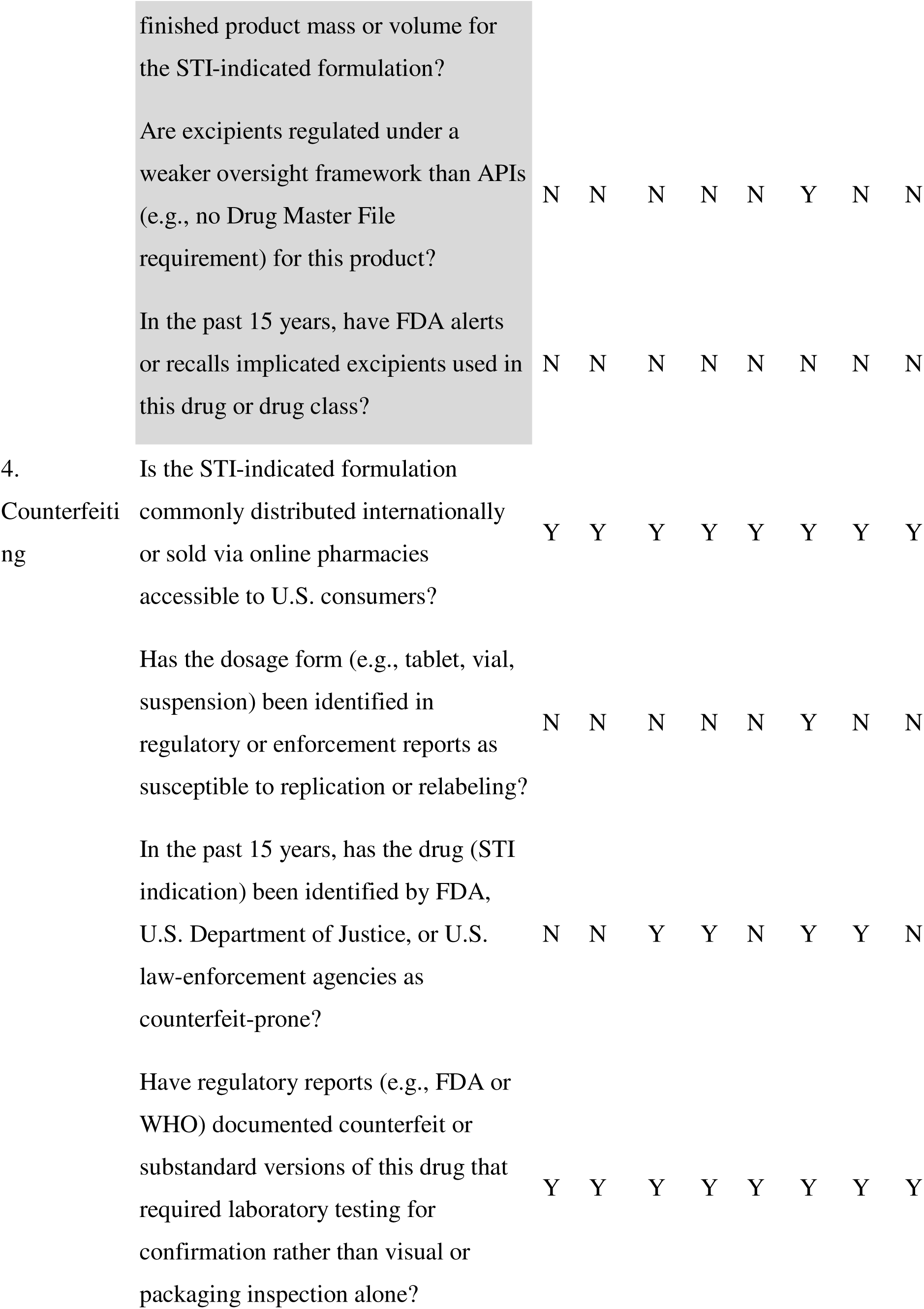
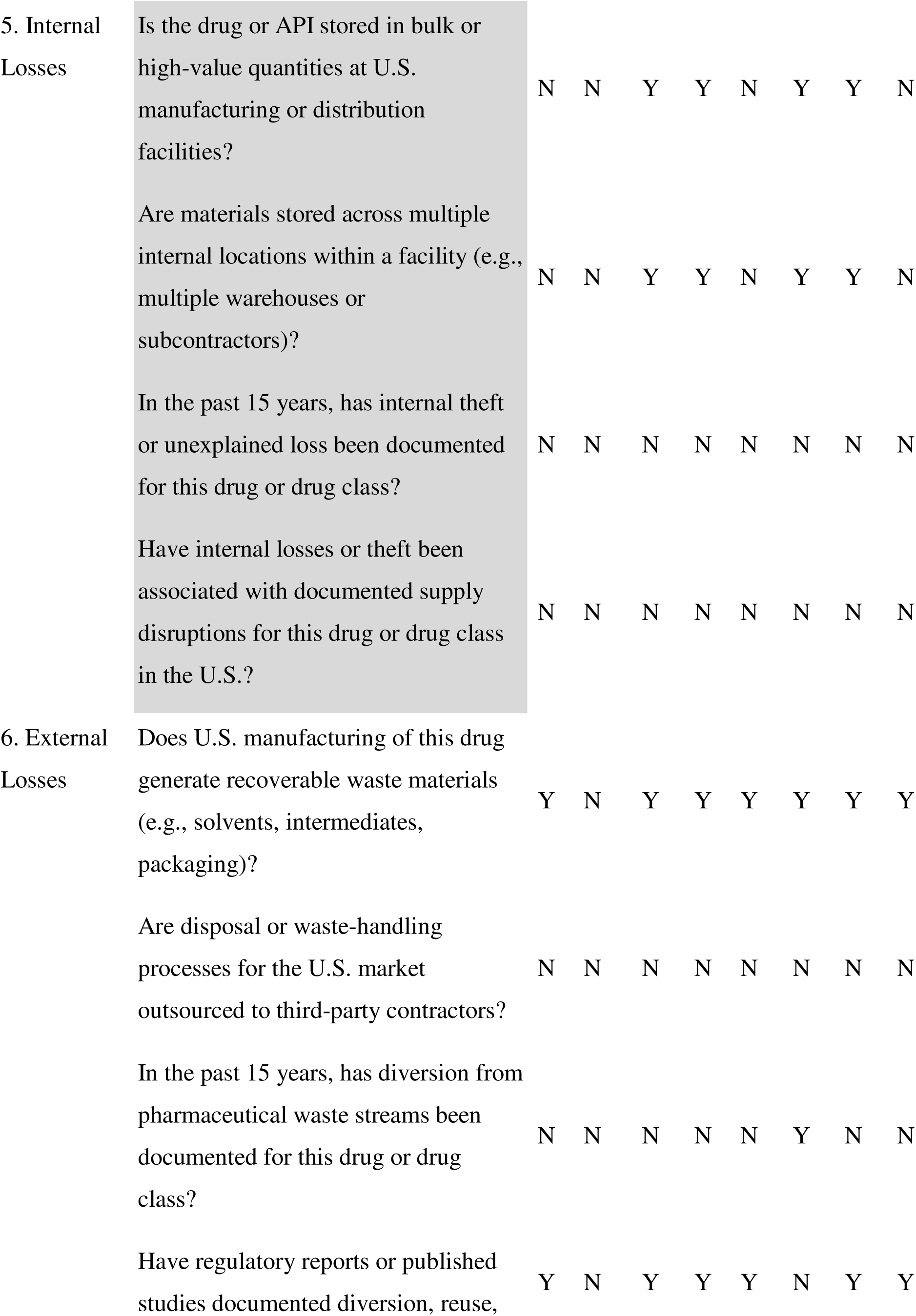
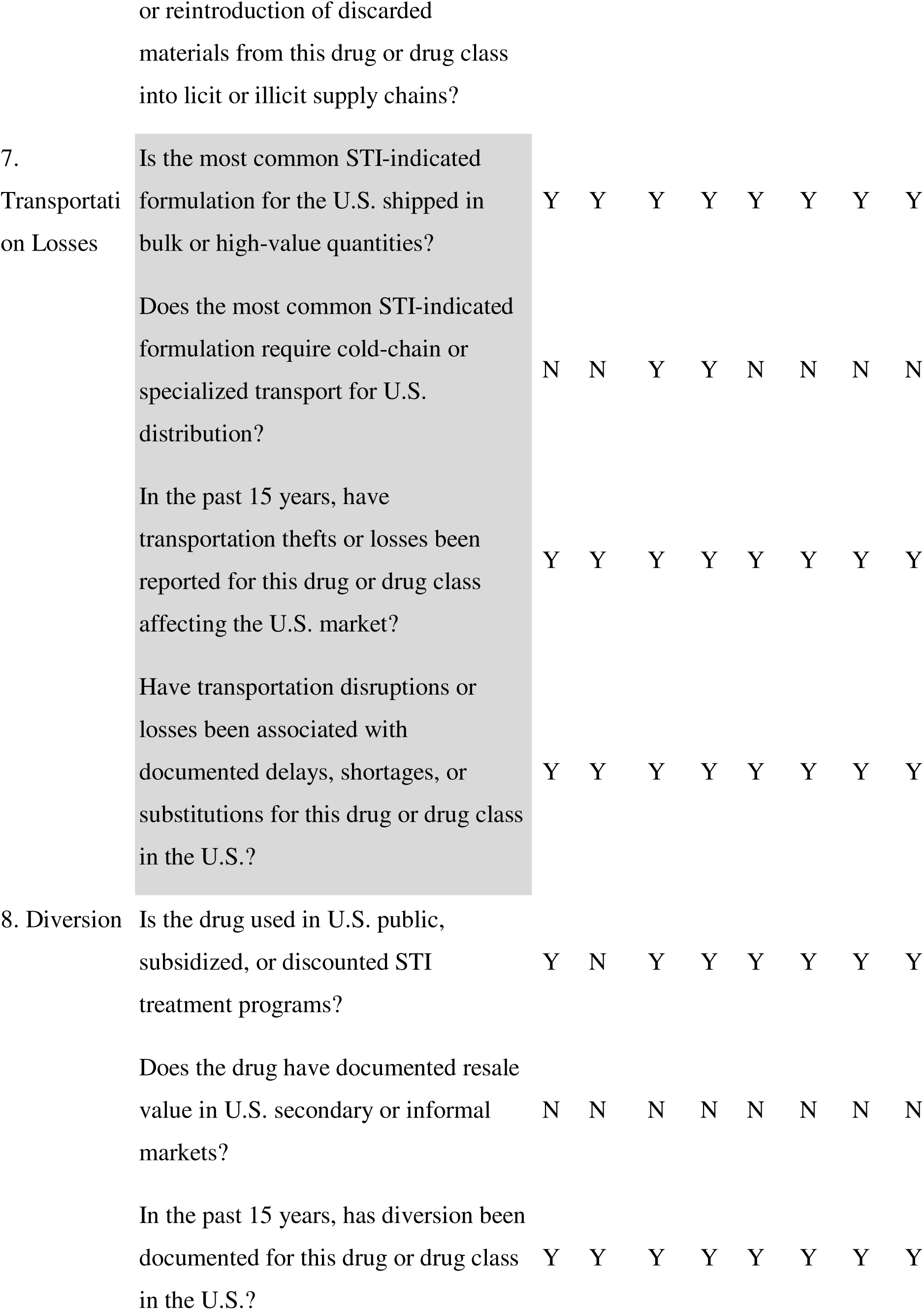
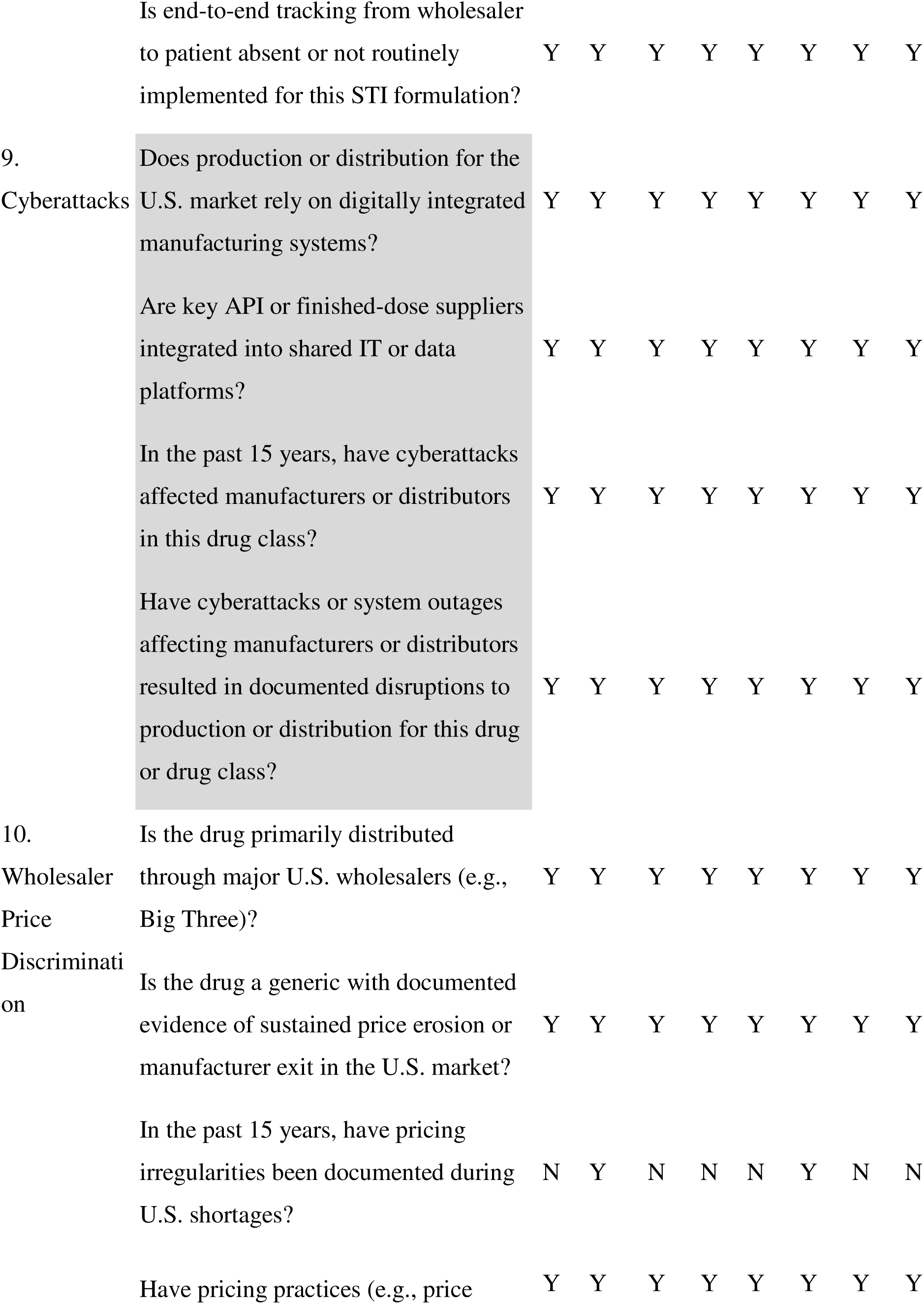
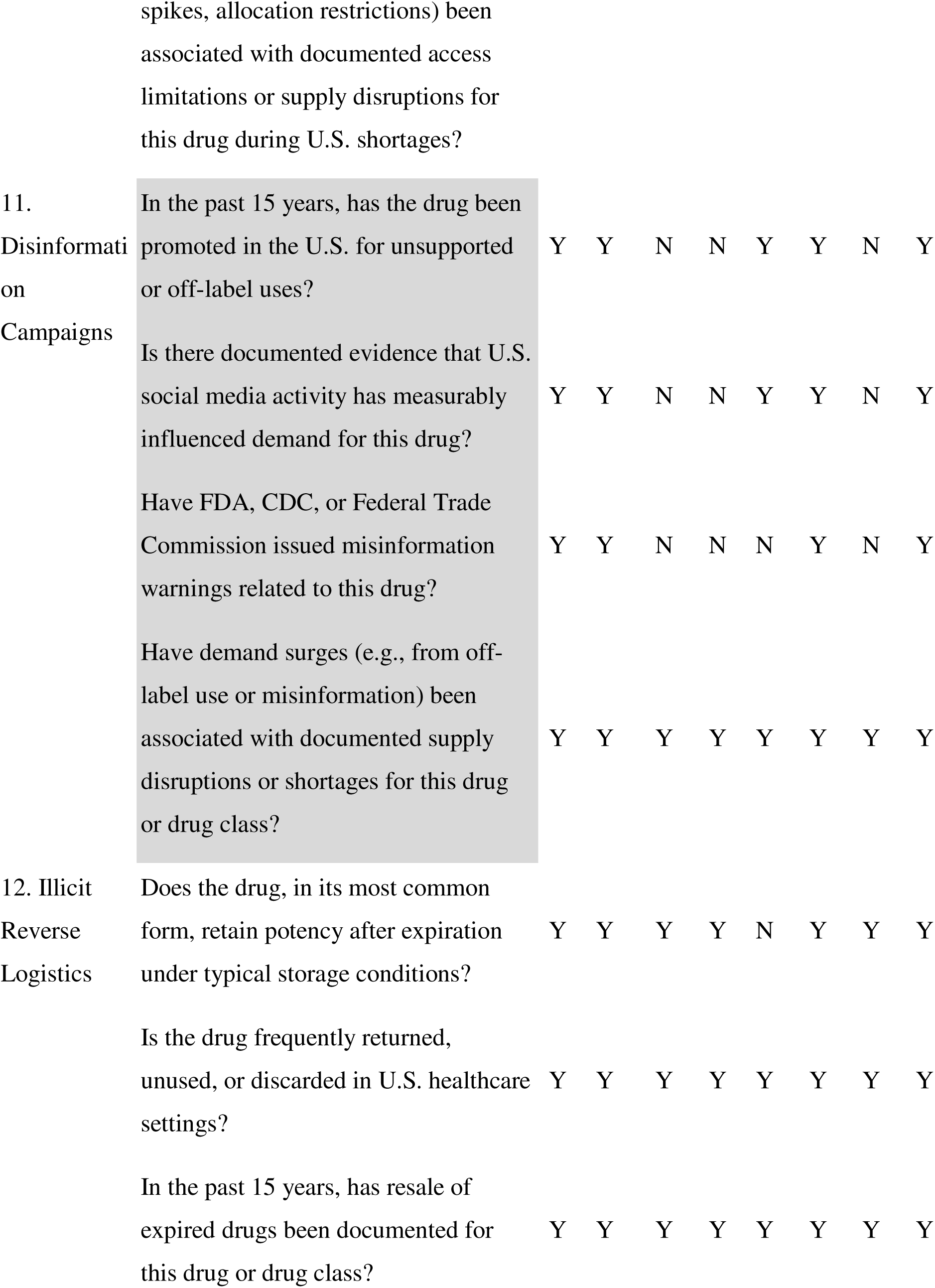
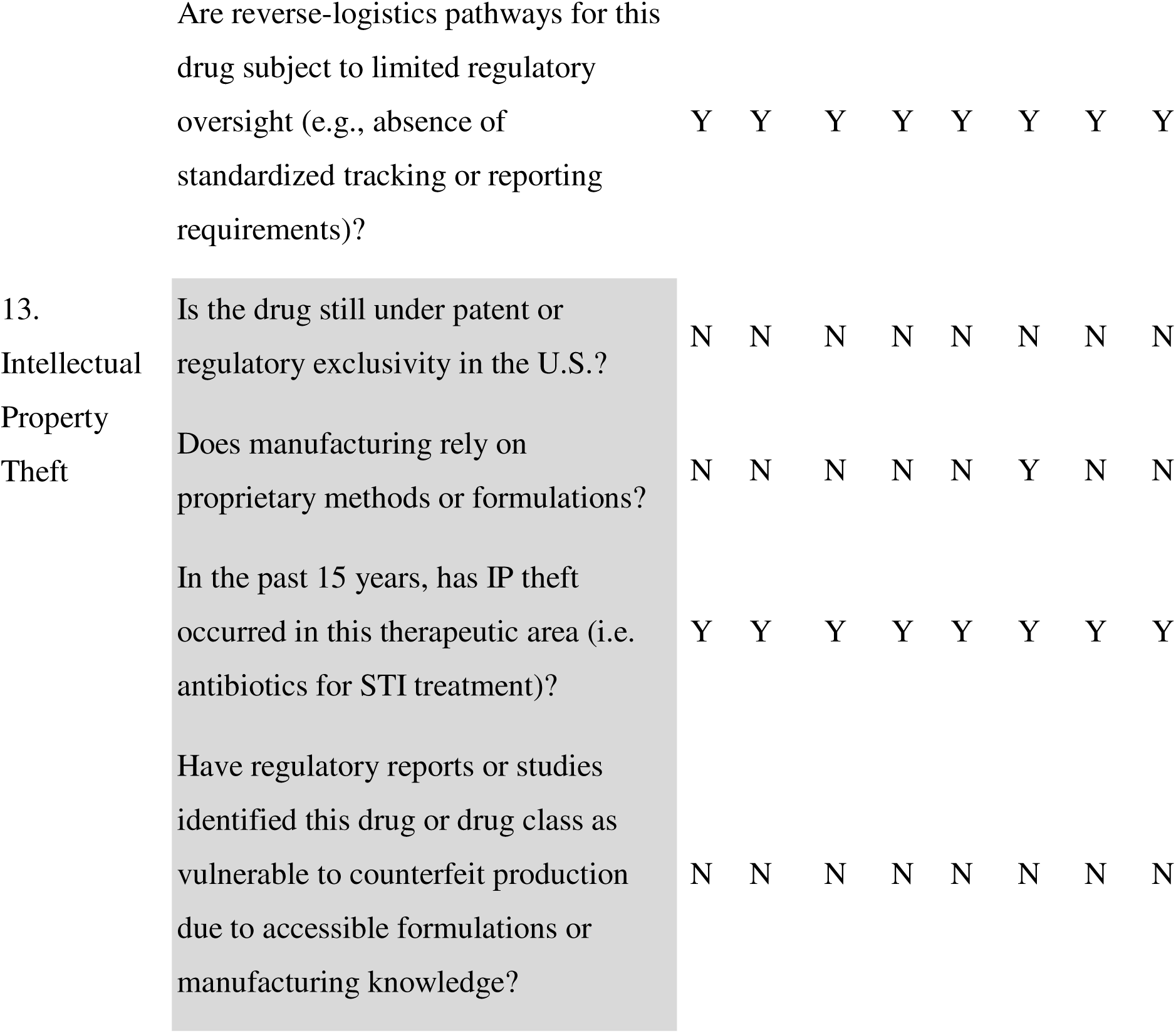
Binary Classification of Supply-Chain Vulnerabilities Across Eight First-Line STI Medications Using a 13-Category Disruption Framework, United States, 2022–2025.

### Supply-Chain Vulnerability Framework

Application of the supply-chain disruption framework identified vulnerabilities across multiple stages of the pharmaceutical supply chain, including active pharmaceutical ingredient sourcing, sterile manufacturing processes, distribution systems, market structure, and demand variability (Table 3).

Sterile injectable drugs, ceftriaxone and benzathine penicillin G, require aseptic manufacturing, sterile fill–finish production, and specialized packaging components such as sterile vials or prefilled syringes. Those manufacturing requirements necessitate additional production stages and quality-control processes not present in oral solid-dose manufacturing. Oral antibiotics including azithromycin, doxycycline, metronidazole, and cefixime relied on globally distributed ingredient manufacturing and multi-stage supplier networks.

All eight drugs relied on globally distributed active pharmaceutical ingredient supply chains, with ≥50% of active pharmaceutical ingredient volume manufactured outside the U.S. or European Union (Table 3). Upstream sourcing networks involving multiple intermediaries (≥3) were identified for all drugs, indicating widespread exposure to multi-step supply dependencies.

Market structure vulnerabilities were observed across the generic drug set. All eight drugs were generic products distributed through major U.S. pharmaceutical wholesalers and characterized by documented evidence of sustained price erosion or manufacturer exit (Table 3). Evidence of pricing irregularities during shortages was identified for azithromycin and doxycycline, while benzathine penicillin G demonstrated extreme supplier concentration (≤3 active pharmaceutical ingredient manufacturers supplying ≥75% of demand).

Demand-side vulnerabilities were also identified. Evidence from regulatory communications, media reports, and prior studies suggested misinformation-related or off-label demand surges was documented for azithromycin, doxycycline, cefixime, and tinidazole, including instances where social media activity promoted unsupported uses and associated with reported increases in utilization, prompting regulatory warnings and, in some cases, contributing to supply disruptions (Table 3).

Across distribution and system-level domains, evidence consistent with vulnerabilities related to transportation disruptions, cyberattacks, and diversion was identified across all drugs, supported by documented incidents such as shipment delays, cyberattacks on pharmaceutical manufacturers and distributors, and diversion within supply chains.

The binary classification matrix summarizing disruption categories for each drug is presented in Table 3. This matrix represents an exploratory application of the framework and does not provide a quantitative ranking of drug-specific vulnerability.

## DISCUSSION

We conducted a structured supply-chain vulnerability assessment of eight first-line therapies used to treat priority STIs in the U.S. . Our analysis identified multiple supply-chain vulnerabilities across those drugs, including internationally concentrated active pharmaceutical ingredient production, manufacturing complexity in sterile injectable formulations, economic pressures within generic drug markets, and emerging disruption pathways, including misinformation-driven demand surges and cybersecurity-related disruptions affecting pharmaceutical distribution systems.

The recent U.S. shortage of benzathine penicillin G demonstrated how structural fragilities in generic pharmaceutical supply chains can rapidly translate into public health consequences ^1^. Prior studies of drug shortages consistently identify manufacturing concentration, thin profit margins, and limited production redundancy as central drivers of supply instability, particularly for sterile injectable generics ^3,19,20^. Our findings suggest that those structural characteristics may extend beyond benzathine penicillin G to multiple first-line STI treatments.

Interruptions in the supply of STI treatments can have direct clinical and public health consequences. Untreated chlamydia and gonorrhea can lead to pelvic inflammatory disease, infertility, ectopic pregnancy, and chronic pelvic pain ^6^. Trichomoniasis, syphilis, and genital herpes are associated with adverse pregnancy and neonatal outcomes including preterm birth, miscarriage, stillbirth, neonatal death, and congenital infection ^6^. All of those STIs have been associated with increased HIV transmission risk.^6^ Reduced availability of effective therapies may force use of less effective or more toxic regimens, lowering cure rates and increasing transmission ^21^. Because STI control depends on rapid diagnosis and treatment, sustained drug availability is critical to prevention infrastructure.

Antibiotic shortages are not new. Penicillin G shortages have been documented since the 1970s and have frequently been associated with manufacturing transitions, facility closures, or limited producer redundancy ^22^. Over time, localized disruptions have become embedded within globally distributed pharmaceutical supply networks. Contemporary shortages increasingly reflect geographically concentrated active pharmaceutical ingredient manufacturing and reliance on a limited number of production facilities, creating single-point failure risks ^22,23^. Our analysis identified reliance on internationally distributed active pharmaceutical ingredient manufacturing across several drugs evaluated in this study. More than one quarter of active pharmaceutical ingredient manufacturing facilities supplying the U.S. market are domestic, while 72% are located overseas, with substantial shares in China (13%) and India (18%) ^24–27^. Outsourcing of active pharmaceutical ingredient production for antibiotics to Asia has intensified that geographic concentration over recent decades ^28,29^.

Trade patterns further highlight the uneven structure of pharmaceutical supply chains. Whereas benzathine penicillin G depends on a highly specialized and fragile production network, often limited to a small number of upstream suppliers and prone to recurring global shortages, azithromycin is produced within a broad, diversified generic market that supports large-scale, reliable distribution. By 2016, just three Chinese active pharmaceutical ingredient manufacturers supplied the global penicillin market, whereas azithromycin was among the most frequently prescribed antibiotics, accounting for 26.3% of local purchases and 73.7% from foreign sources ^30,31^. That contrast illustrates how concentrated production may create persistent supply vulnerabilities, while competitive manufacturing enhances resilience. Adding to this complexity, India remains dependent on Chinese upstream chemical inputs for pharmaceutical ingredients used in the production of many generic antibiotics, including azithromycin; this interdependence introduces additional exposure to bilateral trade disruptions affecting antibiotic supply chains ^32^.

Supply disruptions in concentrated manufacturing systems may propagate rapidly. For example, contamination in a single Italian manufacturing facility combined with raw material shortages in China threatened nearly 60% of Japan’s cefazolin supply ^33^. Limited transparency regarding active pharmaceutical ingredient supplier identity and production capacity further complicates supply-chain monitoring. Manufacturers may rely on intermediaries and therefore lack direct visibility into upstream ingredient sources, reducing the ability of regulators or purchasers to detect emerging supply bottlenecks ^2^. Those structural characteristics specifically affected azithromycin, doxycycline, metronidazole, and cefixime ^34^.

Sterile injectable drugs present additional manufacturing constraints. Ceftriaxone and benzathine penicillin G require aseptic manufacturing, sterile fill–finish production, and specialized packaging components. Those processes are technically complex, capital intensive, and sensitive to quality failures, factors that frequently contribute to drug shortages ^3^.

Ceftriaxone injection products have experienced recalls due to particulate contamination, including rubber particles originating from vial stoppers, illustrating how failures in packaging or aseptic processing can interrupt supply even when active pharmaceutical ingredient availability remains stable ^35^. National analyses indicated that parenteral drugs accounted for 71.7% of shortages in critical care medications over a 15-year period ^36^.

Economic characteristics of generic pharmaceutical markets also contribute to supply instability. Many STI treatments evaluated in this study are long-established generics with limited pricing flexibility and narrow profit margins. An economic analysis found that low profit margins, price volatility, and manufacturer exit from unprofitable therapeutic classes reduced production redundancy and increased the likelihood of shortages ^37^. Procurement systems that emphasize lowest-cost purchasing may further reduce incentives for sustained manufacturing capacity ^38^.

Emerging disruption mechanisms also affect pharmaceutical supply systems. Demand surges driven by misinformation or off-label prescribing can rapidly destabilize supply ^39^. During the COVID-19 pandemic, increased off-label use of azithromycin produced documented supply distortions in several markets ^40^. For example, the azithromycin market in Brazil experienced a profound supply distortion, with consumption surging by 62.8% due to political endorsements and off-label prescribing “COVID kits” ^41^. Cybersecurity incidents pose additional risks; attacks on healthcare logistics systems have disrupted medication ordering and distribution networks in multiple settings ^42^. The WHO estimated that approximately 10% of medical products in low-and middle-income countries are substandard or falsified, with antibiotics among the most affected categories ^43^. Digital traceability systems, including blockchain-based distribution networks, have been proposed to improve supply transparency and reduce counterfeit circulation^44^. Although regulatory oversight is stronger in the U.S., online purchasing channels and international supply networks may still introduce counterfeit or diverted medicines into the market ^45^.

Taken together, our findings indicate that vulnerabilities affecting benzathine penicillin G are not unique to a single drug but arise from broader structural characteristics of generic pharmaceutical supply chains. Supply instability reflects interactions between manufacturing concentration, global ingredient sourcing, market incentives, regulatory oversight, and distribution infrastructure. Addressing those vulnerabilities may require coordinated policy interventions, including diversification of active pharmaceutical ingredient sourcing, increased manufacturing redundancy, improved supply-chain transparency, and economic incentives that support sustainable generic production capacity ^46^. Stronger supply chain integration and coordination are associated with improved resilience under disruption conditions ^10^. Strengthening supply-chain resilience is therefore an important component of maintaining reliable access to essential STI treatments.

From a public health practice perspective, those findings suggest several actionable strategies. First, health systems and public health agencies should monitor supplier concentration for essential antimicrobials and prioritize procurement from diversified manufacturers. Second, policymakers should consider incentives for domestic or geographically distributed active pharmaceutical ingredient production to reduce reliance on concentrated global sources. Third, surveillance systems could incorporate early warning indicators such as demand surges and manufacturer exits. Finally, integrating supply-chain risk assessments into STI control programs may improve preparedness for future disruptions.

### Limitations

This study has several limitations. First, the framework used a binary classification approach rather than quantitative risk scoring. While that permitted consistent comparison across drugs, it precluded estimating the magnitude or probability of disruption. Second, some disruption categories, such as misinformation-driven demand surges and cybersecurity incidents, were not systematically captured in regulatory or bibliographic databases. Evidence for those mechanisms relied partly on gray literature and event reporting, which may be incomplete or uneven across drugs and could lead to under-identification of certain vulnerabilities. Third, our analysis relied on qualitative synthesis of regulatory records, published literature, and documented supply-chain events, which limits causal inference between identified vulnerabilities and observed or future drug shortages. Fourth, the binary classification approach may oversimplify heterogeneity in the severity and likelihood of disruption across drugs and categories. Future research incorporating quantitative market concentration metrics, geographic mapping of active pharmaceutical ingredient production, and longitudinal shortage data could improve risk estimation and modeling of supply-chain disruption.

## Conclusion

Multiple structural vulnerabilities affect eight first-line therapeutics for the five priority sexually transmitted pathogens, including reliance on internationally distributed active pharmaceutical ingredient production, manufacturing constraints associated with sterile injectable formulations, market concentration within generic drug manufacturing, and documented disruption events affecting pharmaceutical supply systems.

## DATA AVAILABILITY

Data supporting this study is available upon request. A detailed spreadsheet containing the data, along with source links, can be accessed by contacting the corresponding author.

## STATEMENTS AND DECLARATION

### Author Contributions

Coby Y. Garcia (CYG) -, Methodology, Investigation, Formal Analysis, Results Interpretation, Writing - Original Draft, Writing - Review & Editing

William Leung (WL) - Data Curation, Investigation, Writing - Review & Editing Amelia Shirley (AS) - Data Curation, Investigation, Writing – Reviewing & Editing

Lao-Tzu Allan-Blitz (LAT) - Conceptualization, Project Administration, Supervision, Results Interpretation, Writing - Review & Editing

## ABREVATIONS

FDA: U.S. Food and Drug Administration
HIV: Human Immunodeficiency Virus
WHO: World Health Organization
CDC: Centers for Disease Control and Prevention
STI: Sexually Transmitted Infection
API: Active Pharmaceutical Ingredient
U.S.: United States
EU: European Union

## Appendix

**Table S1.** Characteristics of Excluded Pharmacologic Agents and Reasons for Exclusion from Supply-Chain Vulnerability Analysis, United States, 2021–2025.

| Drug to Exclude | Primary Reason for Exclusion | Detailed Context |
| --- | --- | --- |
| Valacyclovir | Prodrug/Redundancy | Delivers the same active ingredient as Acyclovir. Exclusion simplifies the list by removing therapeutic duplication. |
| Gentamicin | Niche Alternate/Allergy | Only used for Gonorrhea as a second-line option for patients with severe cephalosporin allergy. |
| Foscarnet | Salvage Therapy/Rare Use | Reserved for rare, drug-resistant Herpes cases, typically in immunocompromised patients; far outside standard treatment. |
| Levofloxacin | Non-Standard Alternate | A Fluoroquinolone antibiotic used as a less-preferred alternative for Chlamydia. |
| Amoxicillin | Niche Use/Pregnancy | Used only as a specific, second-choice alternative for Chlamydia treatment in pregnancy. |
| Famciclovir | Prodrug/Redundancy | Like Valacyclovir, it's an alternative antiviral prodrug for Herpes, which is covered by Acyclovir. |

### Search Strategy, Evidence Sources, and Binary Assessment Framework

1. Bibliographic Database Search Strategy Peer-reviewed evidence was identified through structured searches of MEDLINE (via PubMed) and Embase (via Elsevier Embase interface). Searches were conducted between January 1, 2015 and January 31, 2026. For each drug included in the analytic sample (acyclovir, azithromycin, benzathine penicillin G, cefixime, ceftriaxone, doxycycline, metronidazole, and tinidazole), searches combined the generic drug name with disruption-related terms designed to identify evidence of pharmaceutical supply-chain vulnerabilities. The core search syntax was: ("drug name") AND ("drug shortage" OR "supply chain" OR "manufacturing disruption" OR "manufacturing quality issue" OR "recall" OR "active pharmaceutical ingredient" OR "API shortage" OR "production capacity" OR "supply disruption" OR "distribution disruption" OR "demand surge" OR "counterfeit" OR "diversion" OR "cyberattack" OR "export restriction" OR "trade dispute") Searches were limited to English-language publications. Search results were screened to identify evidence describing all 13 categories.
2. U.S. Regulatory and Market Databases To characterize regulatory status and market structure, we reviewed the following databases:

a. Drugs@FDA: Used to identify approved New Drug Applications (NDAs) and Abbreviated New Drug Applications (ANDAs).
b. FDA Drug Shortages Database: Used to identify (1) active shortages, (2) resolved shortages, (3) manufacturer-reported causes of shortage, and (4) estimated resupply timelines.
c. Approved Drug Products with Therapeutic Equivalence Evaluations (Orange Book): Used to assess: (1) therapeutic equivalence codes, (2) approval history, and (3) application holders.
d. National Drug Code Directory: Used to identify: (1) marketed products, (2) dosage forms, (3) packaging presentations and (4) active labelers. These databases were accessed in December 2025.
3. Gray Literature and Event Detection Strategy Certain disruption categories, such as misinformation-driven demand surges, cyber incidents, and trade disputes, are not traditionally systematically indexed in bibliographic databases. To identify evidence for these domains, we conducted structured gray-literature searches. Searches were performed using the Google search engine and targeted website queries.
  a. U.S. Government Sources: The following domains were searched: The following domains were searched: fda.gov, cdc.gov, hhs.gov, aspr.hhs.gov, dea.gov, cbp.gov.
  b. International and Multilateral Organizations: The following domains were searched: who.int, oecd.org, worldbank.org.
  c. Industry and Supply-Chain Sources: Sources reviewed included manufacturer recall notices, pharmaceutical wholesaler communications, and pharmacy-organization publications when directly relevant to the assessment question.
  d. News and Archival Sources: To identify publicly reported disruption events, we searched: Archival and media sources included Reuters, Bloomberg, major national newspapers, and LexisNexis or similar news databases where available. These sources were used to identify publicly reported events such as manufacturing shutdowns, contamination events, export restrictions, trade disputes, supply disruptions, and cybersecurity incidents affecting pharmaceutical distribution systems.
4. Gray Literature Search Syntax Searches were conducted between January 2024 and January 2026 The core search syntax was: ("drug name") AND ("drug shortage" OR "supply chain" OR "manufacturing disruption" OR "manufacturing quality issue" OR "recall" OR "active pharmaceutical ingredient" OR "API shortage" OR "production capacity" OR "supply disruption" OR "distribution disruption" OR "demand surge" OR "counterfeit" OR "diversion" OR "cyberattack" OR "export restriction" OR "trade dispute")
5. Review Process Three reviewers independently evaluated each drug–disruption category pair using a standardized evidence extraction log. For each binary question, reviewers documented: (1) source, (2) supporting evidence, and (3) classification (yes or no). Disagreements were resolved through discussion and consensus review.

